# Closed environments facilitate secondary transmission of coronavirus disease 2019 (COVID-19)

**DOI:** 10.1101/2020.02.28.20029272

**Authors:** Hiroshi Nishiura, Hitoshi Oshitani, Tetsuro Kobayashi, Tomoya Saito, Tomimasa Sunagawa, Tamano Matsui, Takaji Wakita, MHLW COVID-19 Response Team, Motoi Suzuki

## Abstract

**Objective:** To identify common features of cases with novel coronavirus disease (COVID-19) so as to better understand what factors promote secondary transmission including superspreading events.

**Methods:** A total of 110 cases were examined among eleven clusters and sporadic cases, and investigated who acquired infection from whom. The clusters included four in Tokyo and one each in Aichi, Fukuoka, Hokkaido, Ishikawa, Kanagawa and Wakayama prefectures. The number of secondary cases generated by each primary case was calculated using contact tracing data.

**Results:** Of the 110 cases examined, 27 (24.6%) were primary cases who generated secondary cases. The odds that a primary case transmitted COVID-19 in a closed environment was 18.7 times greater compared to an open-air environment (95% confidence interval [CI]: 6.0, 57.9).

**Conclusions:** It is plausible that closed environments contribute to secondary transmission of COVID-19 and promote superspreading events. Our findings are also consistent with the declining incidence of COVID-19 cases in China, as gathering in closed environments was prohibited in the wake of the rapid spread of the disease.

## Introduction

Although the incidence of coronavirus disease 2019 (COVID-19) in China began to decrease in February 2020,^1^ many countries are struggling with containment of the disease. To effectively reduce the spread of COVID-19, it is vital to identify common features of cases so as to better understand what factors promote superspreading events,^2^ wherein an extraordinarily large number of secondary transmissions are produced by a single primary case. Commissioned by the Minister of the Ministry of Health, Labour, and Welfare of Japan, we collected secondary transmission data with the aim of identifying high risk transmission settings.

## Methods

As of 28 February 2020,^3^ we examined a total of 110 cases among eleven clusters and sporadic cases, and investigated who acquired infection from whom. The clusters included four in Tokyo and one each in Aichi, Fukuoka, Hokkaido, Ishikawa, Kanagawa and Wakayama prefectures. All traced transmission events were examined in relation to close contact in indoor environments, including fitness gyms, a restaurant boat on a river, hospitals, and a snow festival where there were eating spaces in tents with minimal ventilation rate. The number of secondary cases generated by each primary case was calculated using contact tracing data.

## Results

Of the 110 cases examined, 27 (24.6%) were primary cases who generated secondary cases. Figure 1 shows the distribution of these transmissions, of which the mean and variance were 0.6 cases and 2.5 cases^2^, respectively. The odds that a primary case transmitted COVID-19 in a closed environment was 18.7 times greater compared to an open-air environment (95% confidence interval [CI]: 6.0, 57.9).

**Figure 1.**
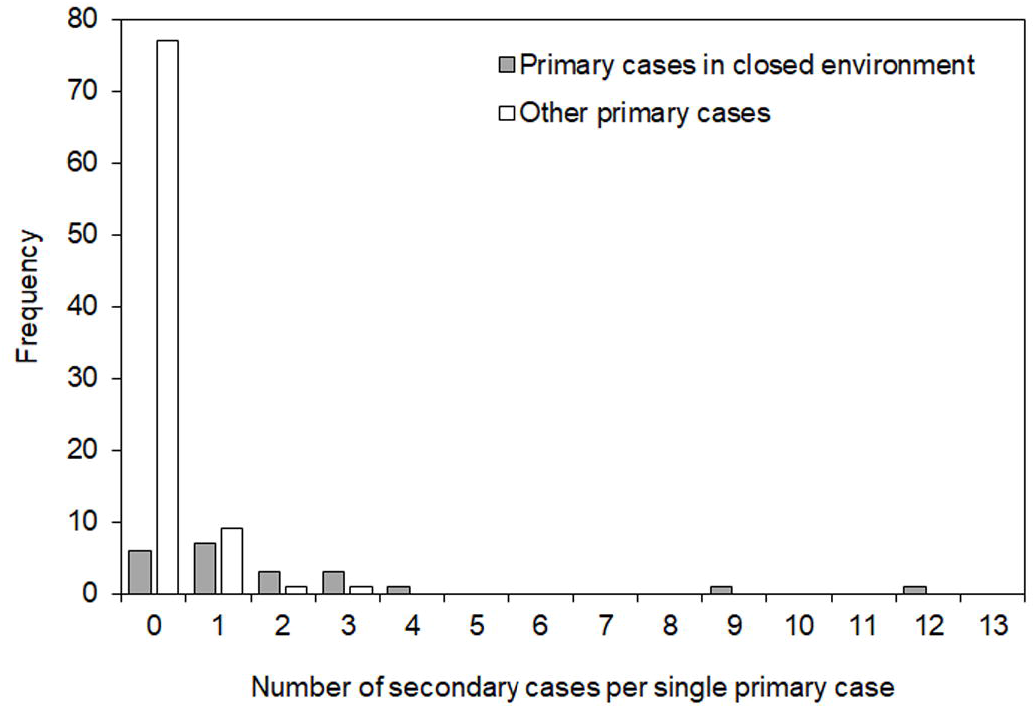
The distribution of the number of secondary cases generated by a single primary case with novel coronavirus (COVID-19). The mean and variance were 0.6 cases and 2.5 cases^2^, respectively.

If superspreading events are defined as events where the number of secondary cases generated by a single primary case is greater than the 95th percentile of the distribution (i.e. transmission to three or more persons), then seven of the 110 cases (6.4%) were involved in such events. Six of these events (85.7%) took place in closed environments, and the odds ratio (OR) of superspreading events in closed environments was as high as 32.6 (95% CI: 3.7, 289.5).

## Discussion

It is plausible that closed environments contribute to secondary transmission of COVID-19 and promote superspreading events. Closed environments are consistent with environmental sampling study^4^ and also large-scale COVID-19 transmission events such as that of the ski chalet-associated cluster in France and the church- and hospital-associated clusters in South Korea^5^. Our findings are also consistent with the declining incidence of COVID-19 cases in China, as gathering in closed environments was prohibited in the wake of the rapid spread of the disease.

Reduction of unnecessary close contact in closed environments may help prevent large case clusters and superspreading events. We hope that with such a reduction in contact the reproduction number of COVID-19 in Japan will be maintained below 1 and contact tracing will be sufficient to contain disease spread.^6^ As the possibility of confounders and interactions was not assessed in this study, additional studies must be conducted to verify the importance of closed environments as facilitators for transmission of COVID-19.

## Data Availability

Anonymized dataset will be provided by the corresponding author upon request.

## Acknowledgement

We sincerely thank staff of local governments, including health centers and prefectural institutes of public health, healthcare facilities, and associated companies and organizations for cooperating us to collect and investigate secondary transmission data. H.N. received funding support from Japan Agency for Medical Research and Development [grant number: JP18fk0108050] and the Japan Science and Technology Agency (JST) Core Research for Evolutional Science and Technology (CREST) program [grant number: JPMJCR1413].

